# Upregulation of ARHGAP9 is correlated with poor prognosis and immune infiltration in clear cell renal cell carcinoma

**DOI:** 10.1101/2023.08.31.23294890

**Authors:** Yu-ling Xiong, Chao Peng, Yue Tian

## Abstract

**Background:** Rho GTPase Activating Protein (ARHGAP) family genes play critical roles in the onset and progression of human cancer. Rho GTPase Activating Protein 9 (ARHGAP9) is upregulated in various tumors. However, far too little attention has been paid to the prognostic value of ARHGAP9 and correlation with immune infiltration in clear cell renal cell carcinoma. Our aim is to evaluate the prognostic significance of ARHGAP9 expression and its correlation with immune infiltration in clear cell renal cell carcinoma.

**Methods:** Transcriptional expression profiles of ARHGAP9 between clear cell renal cell carcinoma tissues and normal tissues were downloaded from the Cancer Genome Atlas (TCGA). The ARHGAP9 protein expression was assessed by the Clinical Proteomic Tumor Analysis Consortium (CPTAC). Receiver operating characteristic (ROC) curve was used to differentiate clear cell renal cell carcinoma from adjacent normal tissues. The Kaplan-Meier method was conducted to assess the effect of ARHGAP9 on survival. Protein-protein interaction (PPI) networks were constructed by the STRING. Functional enrichment analyses were performed using the "ClusterProfiler" package. The immune infiltration patterns were evaluated via the tumor immune estimation resource 2.0 (TIMER 2.0) and TISIDB database.

**Results:** ARHGAP9 expression was substantially higher in clear cell renal cell carcinoma tissues than in adjacent normal tissues. Increased ARHGAP9 mRNA expression was shown to be linked to high TNM stage and lymph node metastases. The diagnostic value of ARHGAP9 gene expression data was assessed using ROC curve analysis. The survival analysis module of GEPIA2 and the Kaplan-Meier plotter both showed clear cell renal cell carcinoma patients with high-ARHGAP9 had a worse prognosis than those with low-ARHGAP9. Correlation analysis indicated ARHGAP9 mRNA expression was significantly correlated with tumor purity and immune infiltrates.

**Conclusion:** These findings demonstrate that up-regulated ARHGAP9 indicates poor prognosis and immune infiltration in clear cell renal cell carcinoma. The current findings suggest that ARHGAP9 can be an effective biomarker and potential therapeutic strategy for ccRCC.

## Introduction

Renal cell carcinoma (RCC) is among the 10 most common cancers worldwide[1], of which clear cell renal cell carcinoma (ccRCC) represents the most common renal cancer histology and accounts for most cancer-related deaths[2]. Around 30% of ccRCC patients have *metastases* at initial presentation[3], the poor prognosis of ccRCC is partly attributable to tumour metastasis[4,5]. Although many therapeutic strategies have been applied to clear cell renal cell carcinoma, advanced ccRCC is a lethal disease, portending a 5-year survival of less than 10%. Clear cell renal cell carcinoma *is reported* highly *resistant* to *chemotherapy* and radiotherapy[6]. Therefore, it is urgent to explore effective therapeutic strategies and novel useful biomarkers for diagnosis and prognosis prediction of ccRCC[7].

Rho GTPase activating protein 9 (ARHGAP9) is a protein coding gene, which encodes a member of the Rho-GAP family of GTPase activating proteins[8,9]. It has been confirmed that Rho GTPases are key regulators of many cell activities, such as cell dynamics, cell growth, intracellular membrane transport, gene transcription, cell cycle progression, and apoptosis[10,11]. Rho GTPases operate as molecular switches which is controlled by GDP–GTP cycle[12,13], and the conversion between the active and inactive form is involved in specifically promoting tumor invasion and metastasis[14,15]. Recent evidence has also shown that dysregulation of Rho GTPase signaling is strongly associated with tumorigenesis and malignant phenotypes, including transformation, cell cycle progression, migration, invasion, metastasis, and drug resistance[16]. Therefore, the more precise function of ARHGAP9 in ccRCC needs to be further investigated.

Increasing studies have demonstrated that the aberrant expression of ARHGAP9 is closely related to the progression of a variety of tumor types, such as breast cancer[17], hepatocellular carcinoma[18], acute myeloid leukemia[19,20], bladder cancer[21]. Thus, ARHGAP9 has great potential to serve as an important biomarker for cancer diagnosis or therapeutic target for multiple cancers. However, no studies have been performed to explore the expression, prognostic values and correlation with immune infiltration of ARHGAP9 in clear cell renal cell carcinoma. To this end, exploring further functions of ARHGAP9 in ccRCC is of great significance in targeted tumor therapy.

In this study, we conducted a systematical analysis of ARHGAP9 expression and patient prognosis using databases including TIMER 2.0, UALCAN, Kaplan-Meier Plotter database, s, as well as investigated the co-expression genes associated with ARHGAP9 expression and discussed their potential functions in clear cell renal cell carcinoma. We further explored the correlation between ARHGAP9 expression and immune cell infiltration of tumours using GEPIA2, TISIDB. Our study provides novel insights into the overexpression of ARHGAP9 and poor survival in ccRCC patients, and provides a potential new molecular therapeutic target for the treatment of ccRCC.

## Materials and Methods

### TCGA Datasets

A total of 611 patients with transcriptional expression data of ARHGAP9 and corresponding clinical information were downloaded from the TCGA official website, which contains 72 normal types and 539 tumor types. The samples selected contained ARHGAP9 gene expression data and associated clinical information, including age, gender, T stage, N stage, M stage, Histologic grade, Pathologic stage.

### Tumor Immune Estimation Resource 2.0 (TIMER 2.0) Database

TIMER2.0 is a comprehensive resource for analyzing gene expression across various types of cancer and immune infiltrates. We performed TIMER2.0 database to draw expression difference of ARHGAP9 using TCGA pan-cancer data and determine the correlation between ARHGAP9 expression of clear cell renal cell carcinoma tissues and six immune cell infiltrations including B cells, CD4 + T cells, CD8 + T cells, macrophages, neutrophils, and dendritic cells (DC).

### Clinical Proteomic Tumor Analysis Consortium (CPTAC) and UALCAN

UALCAN provides a protein expression analysis option using data from the Clinical Proteomic Tumor Analysis Consortium (CPTAC). The CPTAC module of UALCAN was used to analyze the expression levels of ARHGAP9 protein expressions between normal tissues and clear cell renal cell carcinoma tissues.

### Protein-Protein Interaction (PPI) Networks and Functional Enrichment Analysis

The database STRING is a precomputed global resource for the exploration and analysis of the retrieval of interacting genes. We conducted STRING to search for co-expression genes. Protein–protein interaction (PPI) networks were constructed using the STRING database with an interaction score >0.4. Gene ontology (GO) enrichment and Kyoto Encyclopedia of Genes and Genomes (KEGG) pathway analyses of co-expression genes were assessed by the “ClusterProfiler” package.

### Tumor-Immune System Interaction Database (TISIDB)

TISIDB is a website for gene and tumor-immune interactions. In this study, we performed TISIDB to investigate correlations between ARHGAP9 expression and tumor-infiltrating lymphocytes (TILs). The correlations between ARHGAP9 and TILs were analyzed with Spearman’s correlation coefficient test.

### Expression Profiling and Interactive Analysis (GEPIA2) database

GEPIA2 is a valuable and highly cited resource for gene expression analysis based on tumor and normal samples from the TCGA and the GTEx databases. The GEPIA2 survival analysis tool was used to assess the relationship between ARHGAP9 mRNA expression and clear cell renal cell carcinoma prognosis. The calculation of hazard ratios was based on the Cox PH (Proportional Hazards) Model. A P-value ≤0.05 was considered statistically significant.

### Kaplan-Meier Plotter database

Kaplan–Meier-plotter is an online tool to evaluate the effect of gene expression on patient survival. In the study, we used the Kaplan–Meier-plotter database to analyze the correlation between ARHGAP9 mRNA expression and overall survival in clear cell renal cell carcinoma patients.

### Statistical analysis

All statistical analyses were performed with the statistical software R (V 3.6.3) and plotting was performed using the R package ggplot2.

## Results

### Expression levels of ARHGAP9 in pan-cancer analysis

ARHGAP9 gene expression was retrieved using the TIMER2.0 database to determine differences between tumor and normal tissues in various cancers. The result showed that, compared with normal tissues, ARHGAP9 was significantly upregulated in 9 of all 23 cancer types. This data indicated the mRNA expression of ARHGAP9 was abnormally expressed across different cancer types (Fig 1).

**Fig 1.**
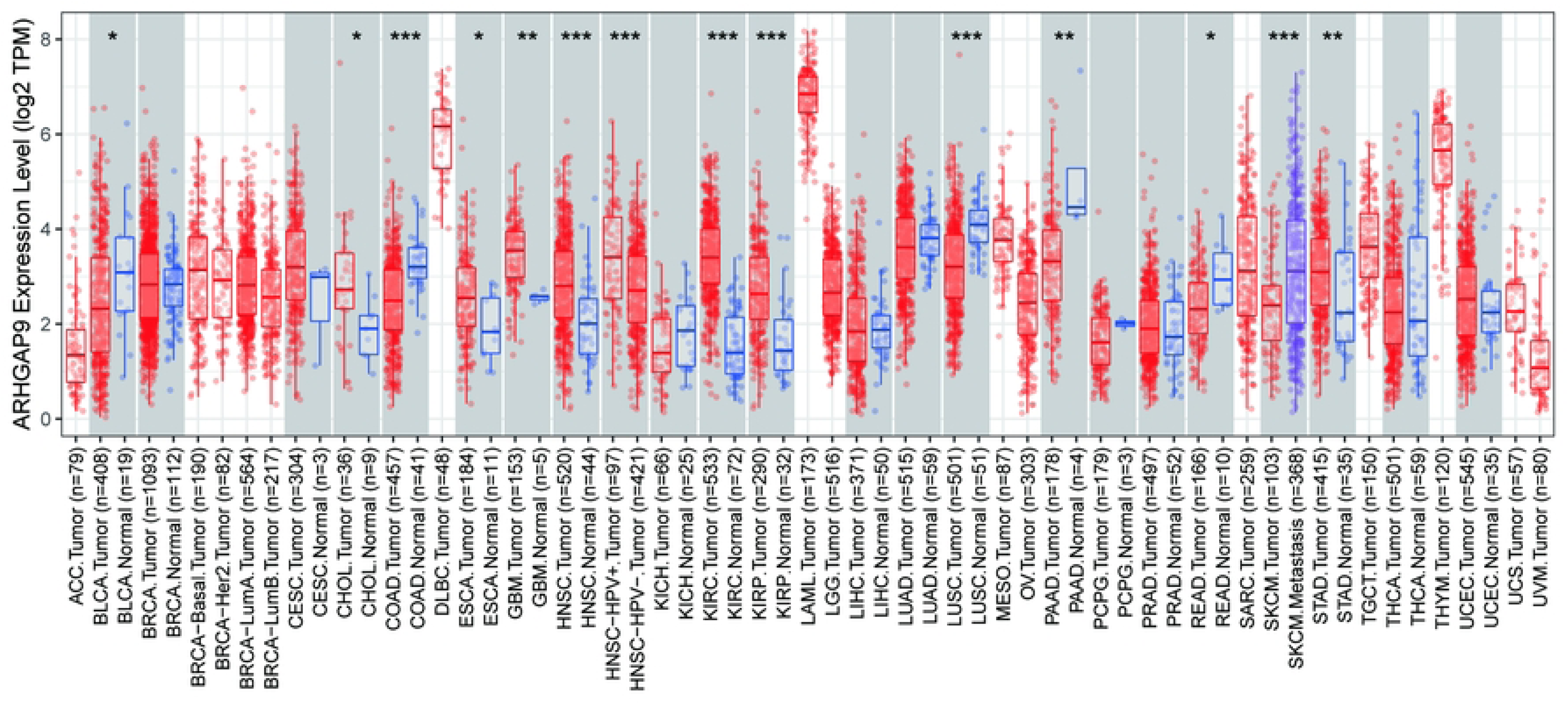
Pan-cancer analysis of ARHGAP9 expression. Human ARHGAP9 expression levels in different cancer types from TCGA data in TIMER2.0. * P < 0.05, **P < 0.01, **P < 0.001. Red indicates tumor tissue and blue indicates normal tissue.

### Upregulated mRNA and protein expression of ARHGAP9 in clear cell renal cell carcinoma patients

ARHGAP9 was validated in protein and mRNA levels using TCGA and the HPA database. As shown in Fig 2A, Paired analysis showed that expression levels of ARHGAP9 were significantly increased in clear cell renal cell carcinoma tissues (n = 72) compared with adjacent non-tumor tissues (n = 72) (Fig 2A, 2.907±0.819 vs. 1.147±0.595, P < 0.001). Unpaired analyses showed that the mRNA expression levels of ARHGAP9 in clear cell renal cell carcinoma tissues (n = 539) were significantly higher than those in adjacent normal tissues (n = 72) (Fig 2B, 2.985 ± 0.918vs. 1.147 ± 0.595, Mann-Whitney U-test, P < 0.001). Analysis of protein expression levels of ARHGAP9 in ccRCC was performed by UALCAN based on the CPTAC database. According to the results, the protein expression of LIMK1 in ccRCC was markedly increased compared to that in normal tissues (Fig 2C). The results revealed that both the mRNA and protein expression of ARHGAP9 are upregulated in clear cell renal cell carcinoma tissues.

**Fig 2.**
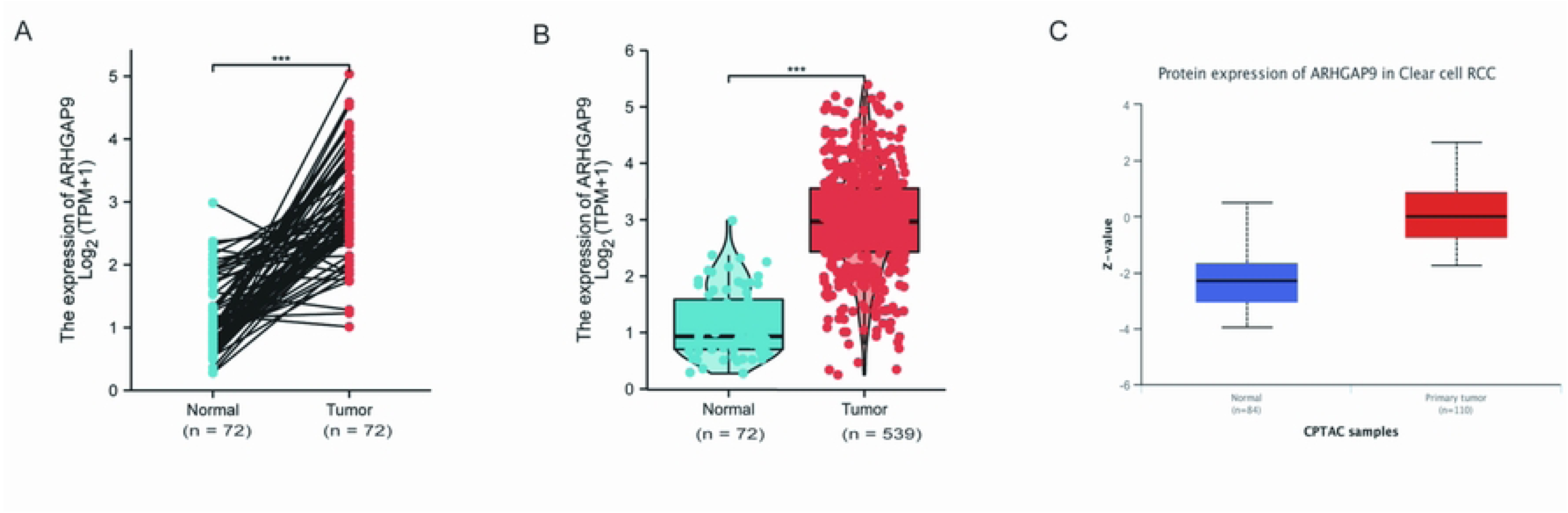
The mRNA expression and protein expression of ARHGAP9 in clear cell renal cell carcinoma. (A) The paird analysis of ARHGAP9 mRNA expression in 72 clear cell renal cell carcinoma samples and matched-adjacent normal samples. (B) The unpaird analysis of ARHGAP9 mRNA expression in 539 l clear cell renal cell carcinoma samples and 72 normal samples. (C) The protein expression levels of ARHGAP9 based on CPTAC.

### Association between ARHGAP9 expression and the clinical pathological characteristics of patients with ccRCC

According to the results (Table 1 and Fig 3A-I), high ARHGAP9 expression was significantly associated with T stage (P = 8.7e−04), N stage (P = 0.02), M stage (P = 1.4e−03), pathologic stage (P = 1.8e−04), and histologic grade (P = 1.6e−04). However, there were no significant associations between ARHGAP9 expression and primary therapy outcome, gender, race, or age (P >0.05). These results indicate that upregulated expression of ARHGAP9 correlates with advanced clinicopathological parameters.

**Fig 3.**
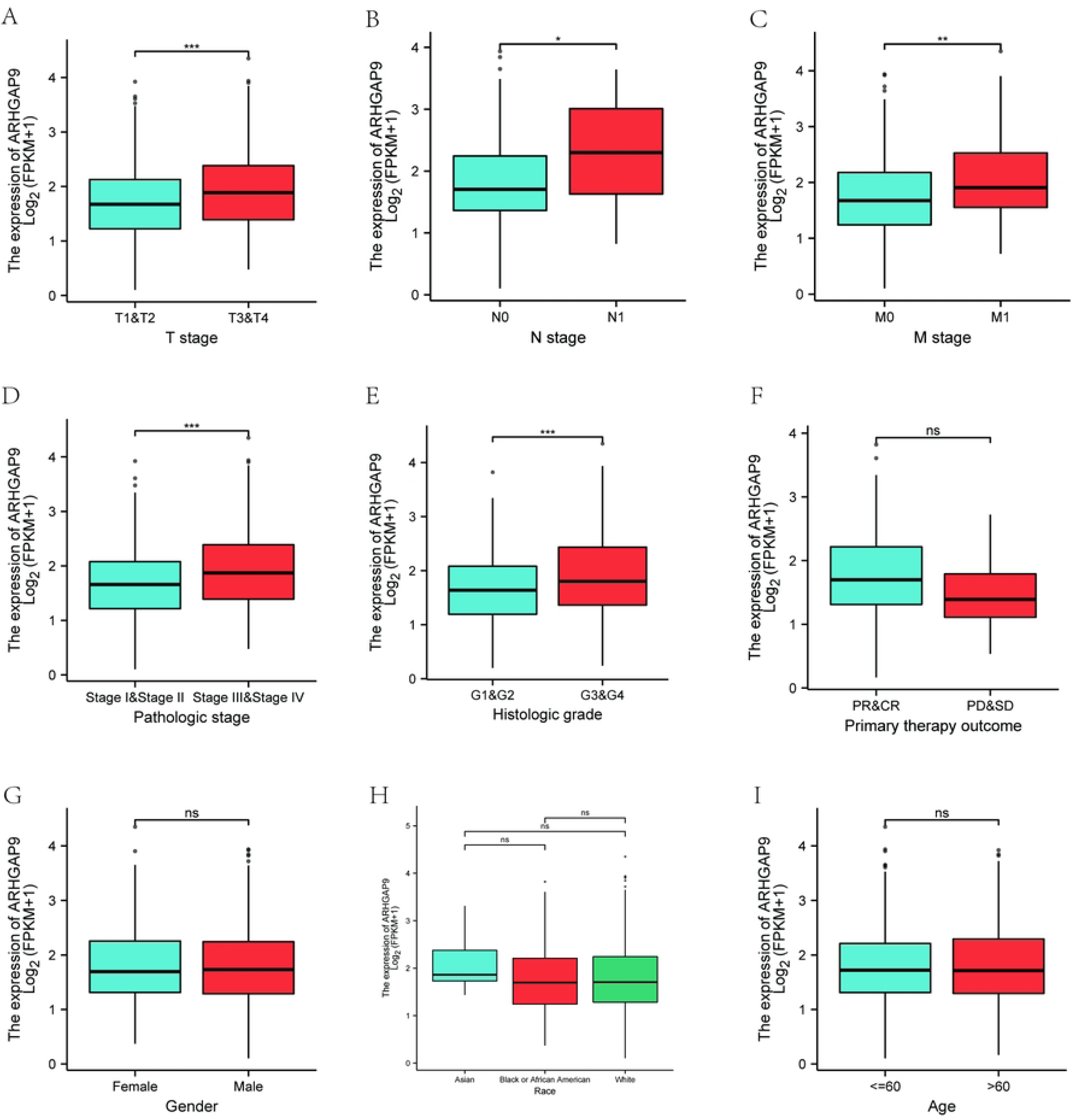
Relationship between ARHGAP9 expression and the clinical pathological characteristics of patients with ccRCC. The correlation between ARHGAP9 expression and clinicopathological characteristics in ccRCC patients, (A) T stage, (B) N stage, (C) M stage, (D) Pathologic stage (P = 0.009), (E) Histologic grade primary therapy outcome, (F) gender, (G) race, and (I) age. *, *P* < *0.05*; **, *P* < 0.01; ***, *P* < *0.001*; ****, *P* < *0.001*; *ns*, nonsignificant.

**Table 1.** Clinical features of the ccRCC (TCGA).

| Characteristic | Low expression of ARHGAP9 | High expression of ARHGAP9 | p |
| --- | --- | --- | --- |
| n | 269 | 270 |  |
| T stage, n (%) |  |  | 0.016 |
| T1 | 157 (29.1%) | 121 (22.4%) |  |
| T2 | 29 (5.4%) | 42 (7.8%) |  |
| T3 | 79 (14.7%) | 100 (18.6%) |  |
| T4 | 4 (0.7%) | 7 (1.3%) |  |
| N stage, n (%) |  |  | 0.214 |
| N0 | 122 (47.5%) | 119 (46.3%) |  |
| N1 | 5 (1.9%) | 11 (4.3%) |  |
| M stage, n (%) |  |  | 0.014 |
| M0 | 227 (44.9%) | 201 (39.7%) |  |
| M1 | 29 (5.7%) | 49 (9.7%) |  |
| Pathologic stage, n (%) |  |  | 0.009 |
| Stage I | 155 (28.9%) | 117 (21.8%) |  |
| Stage II | 25 (4.7%) | 34 (6.3%) |  |
| Stage III | 56 (10.4%) | 67 (12.5%) |  |
| Stage IV | 32 (6%) | 50 (9.3%) |  |
| Primary therapy outcome, n (%) |  |  | 0.072 |
| PD | 5 (3.4%) | 6 (4.1%) |  |
| SD | 6 (4.1%) | 0 (0%) |  |
| PR | 1 (0.7%) | 1 (0.7%) |  |
| CR | 65 (44.2%) | 63 (42.9%) |  |
| Gender, n (%) |  |  | 0.506 |
| Female | 97 (18%) | 89 (16.5%) |  |
| Male | 172 (31.9%) | 181 (33.6%) |  |
| Race, n (%) |  |  | 0.116 |
| Asian | 1 (0.2%) | 7 (1.3%) |  |
| Black or African American | 29 (5.5%) | 28 (5.3%) |  |
| White | 237 (44.5%) | 230 (43.2%) |  |
| Age, n (%) |  |  | 0.897 |
| <=60 | 133 (24.7%) | 136 (25.2%) |  |
| >60 | 136 (25.2%) | 134 (24.9%) |  |
| Histologic grade, n (%) |  |  | 0.002 |
| G1 | 7 (1.3%) | 7 (1.3%) |  |
| G2 | 134 (25.2%) | 101 (19%) |  |
| G3 | 97 (18.3%) | 110 (20.7%) |  |
| G4 | 24 (4.5%) | 51 (9.6%) |  |
| Age, mean ± SD | 60.71 ± 11.99 | 60.54 ± 12.22 | 0.874 |

### Diagnostic value of the ARHGAP9 expression in ccRCC

Receiver operating characteristics (ROC) analysis was used to evaluate the predictive value of the ARHGAP9 expression level. As shown in Fig 4A. The results of ROC analysis revealed that ARHGAP9 expression (AUC: 0.950) had high diagnostic value. Subgroup analysis revealed the predictive value of T I/II (AUC values: 0.940), T III/IV (AUC values: 0.970), N0 (AUC values: 0.956), M1 (AUC values: 0.977), stage I/II (AUC values: 0.938), stage III/IV (AUC values: 0.970), G 1/2 (AUC values: 0.941), G 3/4 (AUC values: 0.969) (Fig 4B–I). These results demonstrated the potential of ARHGAP9 as a diagnostic marker of ccRCC.

**Fig 4.**
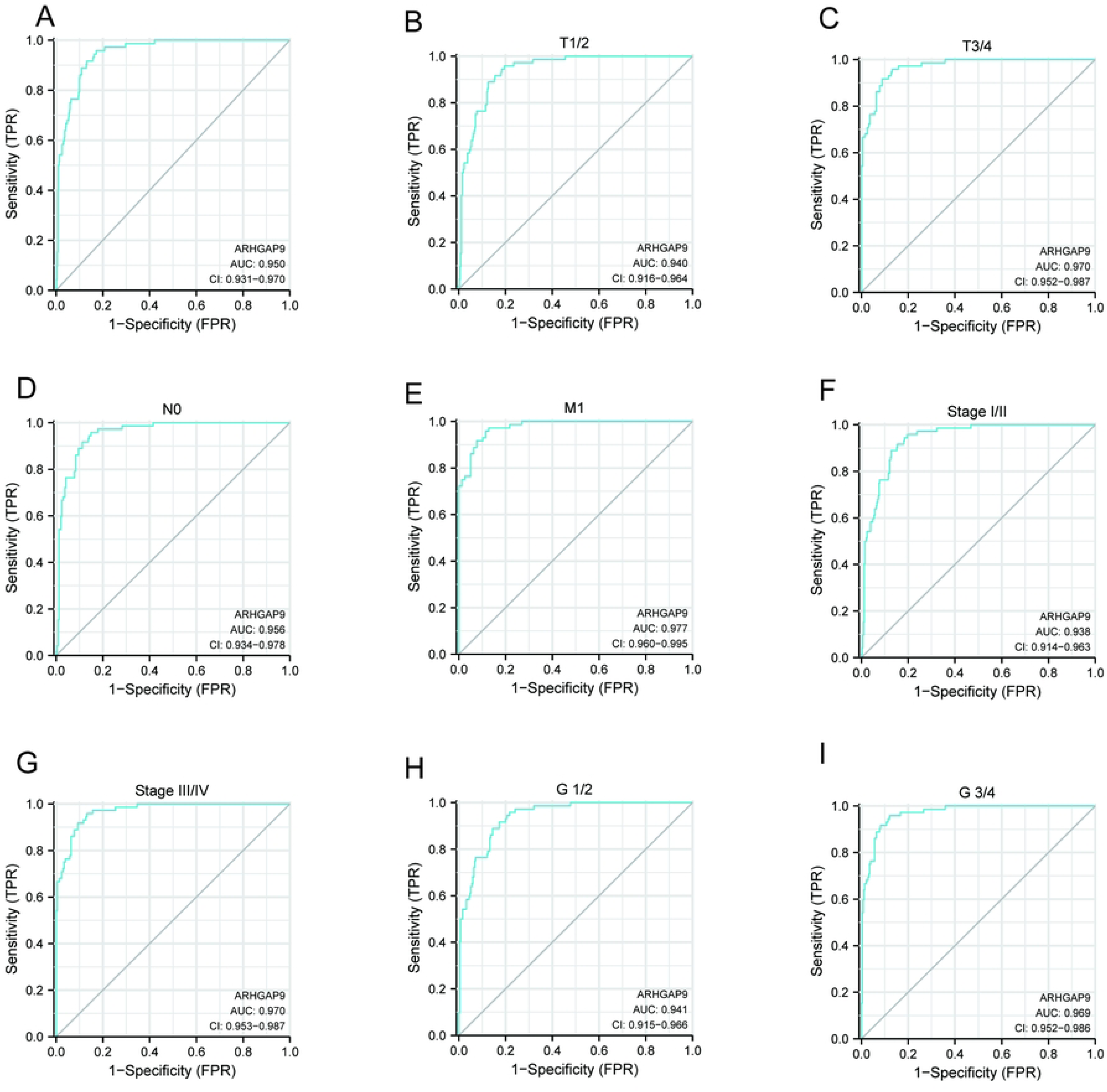
Diagnostic value of ARHGAP9 mRNA expression in ccRCC patients. (A) ROC curve for ARHGAP9 in ccRCC and healthy controls. (B-I) ROC subgroup analysis for T1/2, T3/4, N0, M1, stage I/II, stage III/IV, G1/2, G3/4.

### High expression of ARHGAP9 is a risk factor for short overall survival

To explore the relationship between ARHGAP9 expression and OS in ccRCC patients, the Kaplan-Meier Plotter and the TIMER2.0 database were performed. In Kaplan - Meier Plotter, the OS of clear cell renal cell carcinoma patients with high-level of ARHGAP9 was significantly shorter than those with low-level of ARHGAP9 (OS: HR = 1.83, log-rank P = 0.00027), the results are shown in Fig 5A. The results of TIMER2.0 also indicated that high ARHGAP9 mRNA expression was significantly correlated with short OS in ccRCC patients (OS: HR = 1.5, Cox P = 0.015) (Fig 5B).

**Fig 5.**
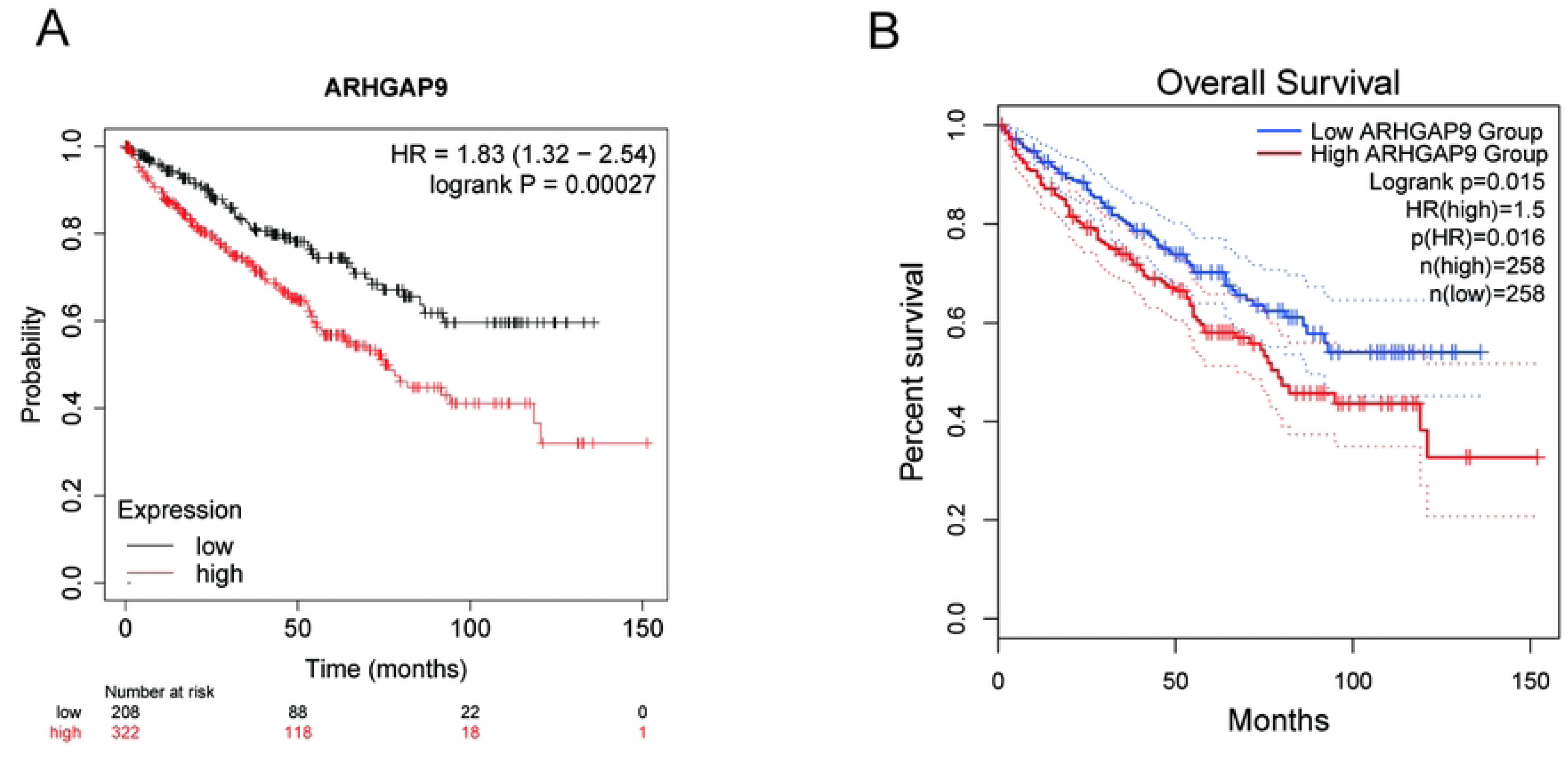
Overall survival analysis of ARHGAP9 mRNA expression. (A, B) The Kaplan-Meier Plotter and TIMER2.0 indicated that patients with high ARHGAP9 expression have a worse prognosis than those with low levels of ARHGAP9.

### Functional annotation and predicted signaling pathways

The PPI network and functional annotations of ARHGAP9 were constructed. The network of ARHGAP9 and its 10 co-expression genes (Rho GTPase-activating protein 30, Epidermal growth factor receptor kinase substrate 8, Growth factor receptor-bound protein 14, Neutrophil cytosol factor 4, Pleckstrin, Ras-related C3 botulinum toxin substrate 1, Ras-related C3 botulinum toxin substrate 2, Transforming protein RhoA, Sphingosine 1-phosphate receptor 4, T-lymphoma invasion and metastasis-inducing protein 1) was visualized (Fig 6A).As shown in Fig 6B, the changes in GO-biological process (GO-BP) of significant genes were mostly enriched for Rho protein signal transduction, Rac protein signal transduction, regulation of small GTPase mediated signal transduction. The changes in GO-cellular component (GO-CC) were significantly enriched for ruffle, eading edge membrane, ruffle membrane. The changes in GO-molecular function (GO-MF) were mainly enriched for phosphatidylinositol binding, phosphatidylinositol phosphate binding, Rac guanyl−nucleotide exchange factor activity. The results of the KEGG pathway analysis indicated that the genes were involved in the Chemokine signaling pathway, the Sphingolipid signaling pathway, the Leukocyte transendothelial migration. As can be seen from Fig 6C–H, the correlation between expression of ARHGAP9 and co-expressed genes in ccRCC patients was analyzed using Pearson correlation analysis.

**Fig 6.**
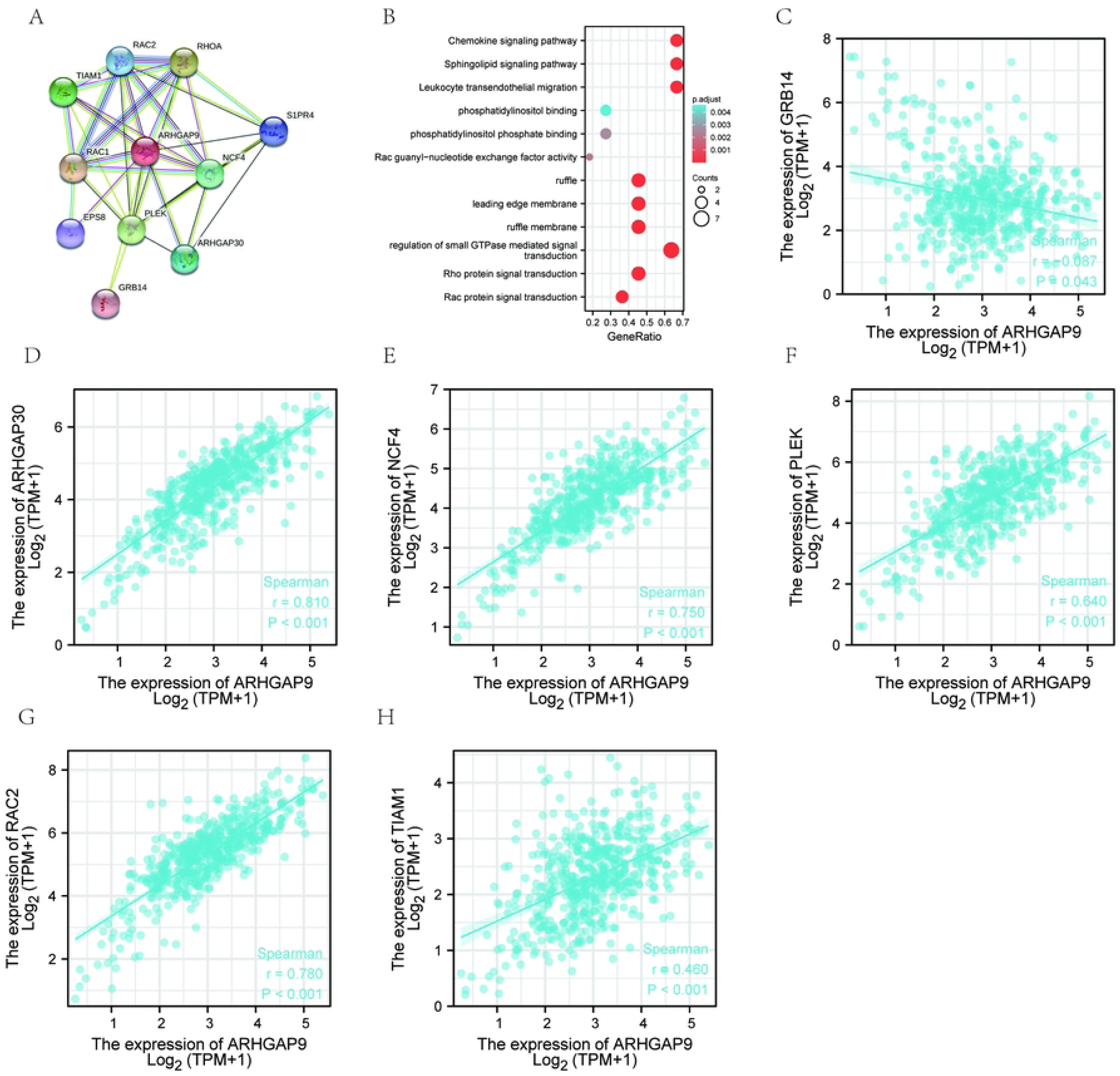
PPI network and functional analysis of ARHGAP9. (A)PPI network of ARHGAP9 and top 10 co-expression genes. (B) Functional enrichment analyses of 11 genes. (C-I) Correlation analysis of ARHGAP9 expression and ten co-expressed genes in ccRCC.

### Immune infiltration level analysis in clear cell renal cell carcinoma associated with ARHGAP9

We performed a correlation analysis between ARHGAP9 gene expression and immune infiltration level in the TIMER database. It can be seen from the data in Fig 7A that ARHGAP9 has a significant correlation with tumor purity in clear cell renal cell carcinoma (r = −0.327, p = 5.24e−13). Accordingly, the ARHGAP9 expression level has significant positive correlations with the infiltration levels of B cells (r = 0.382, p = 2.12e−17), CD8+ T cells (r = 0.475, p = 4.88e−26), CD4+ T cells (r = 0.461, p = 1.29e−25), macrophage cells (r = 0.375, p = 2.30e−16, neutrophils (r = 0.593, p = 7.76e−45), and DCs (r = 0.591, p = 3.02e−44). Fig 7B showed the relationships between the expression of ARHGAP9 and 28 types of TILs across human cancers in the TISIDB database. As shown in Fig 7C, the expression of ARHGAP9 was strongly related to abundance of activated B cells (r = 0.749, p < 0.001), activated CD8+ T cells (r = 0.667, p<0.001), activated CD4+ T cells (r = 0.593, p < 0.001), myeloid derived suppressor cells (MDSC, r = 0.765, p < 0.001), effector memory T cells (Tem CD8, r = 0.74, p < 0.001), immature B cells (r = 0.709, p < 0.001).Our findings suggested that ARHGAP9 may play a specific role in immune infiltration in ccRCC.

**Fig 7.**
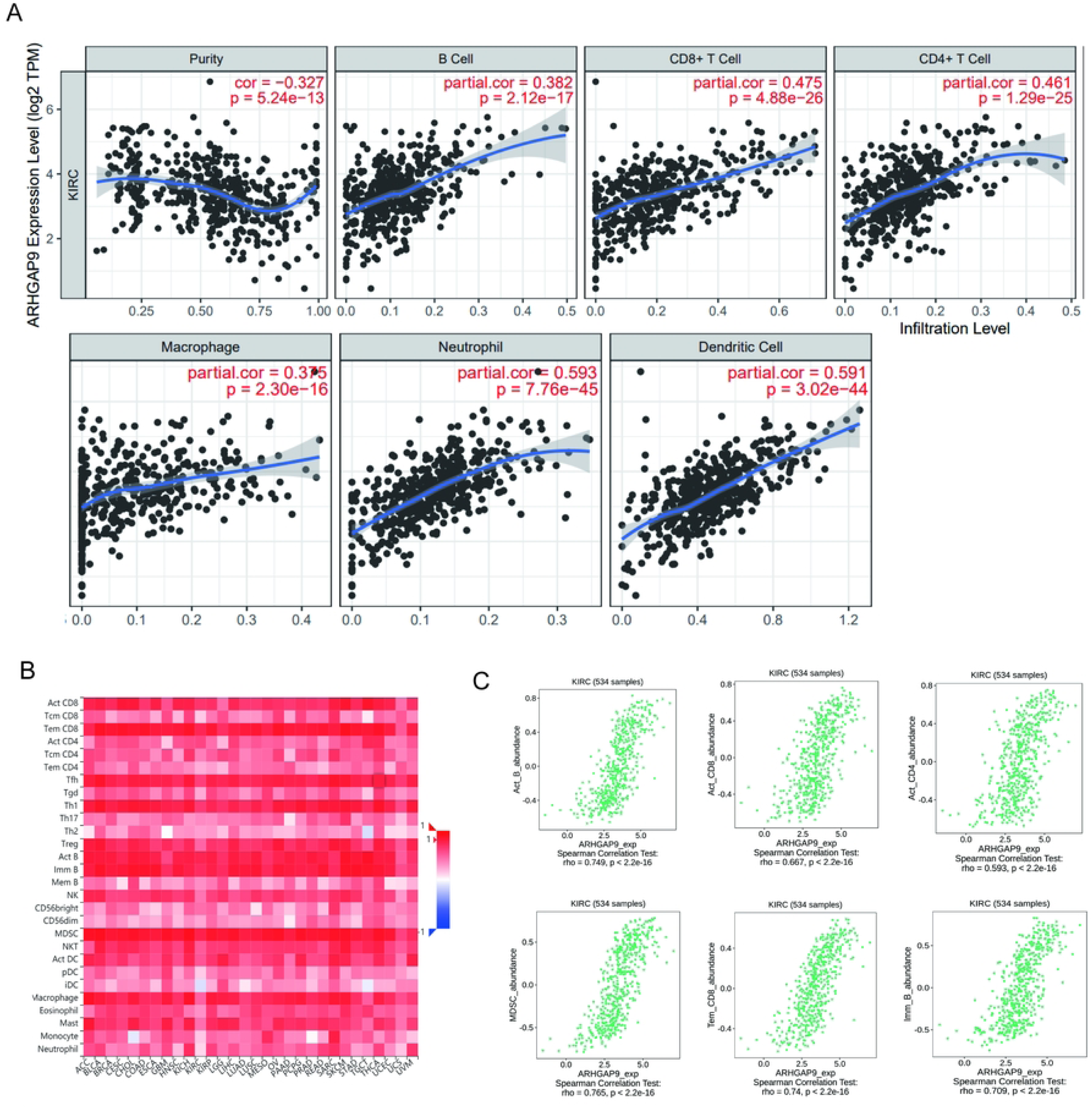
*Correlation between ARHGAP9 expression and immune cell infiltration. (A) Correlations between ARHGAP9 expression and immune infiltration in ccRCC*. (B) correlation analysis between the expression of ARHGAP9 and 28 types of TILs across pan-cancers by TISIDB. (C) ARHGAP9 was related to the abundance of activated B cells, activated CD8+ T cells, activated CD4+ T cells, myeloid derived suppressor cells, effector memory T cells, and immature B cells.

## Discussion

RhoGAP family such as ARHGAP5[22], ARHGAP6[23], ARHGAP7[24], has been reported dysregulated in various cancers, however, prior to this study, the expression and prognostic significance of ARHGAP9 in ccRCC had not been investigated. In this report, for the first time, we found the mRNA expression of ARHGAP9 was higher in clear cell renal cell carcinoma tissues than in normal tissues. The upregulated ARHGAP9 expression is significantly positive-related to advanced TNM stage and lymph node metastases. ROC curve analysis suggested that ARHGAP9 could be a potential diagnostic biomarker to differentiate ccRCC from normal tissues. Kaplan-Meier curves confirmed that high mRNA expression of ARHGAP9 is associated with poor overall survival. Further analysis found that ARHGAP9 was positively correlated with immune infiltration in ccRCC. Therefore, ARHGAP9 may be a valuable prognostic biomarker in patients with ccRCC.

Previous research has shown that ARHGAP9 plays distinct roles in a variety of physiological conditions, tissues, and cells[25]. The up-regulated ARHGAP9 suppressed cell migration and invasion in hepatocellular carcinoma by increasing FOXJ2 expression[18]. In their report, Piao *et al*. reported that a high expression of *ARHGAP9* was significantly associated with prolonged recurrence-free survival in bladder cancer patients[21]. The above reports suggested that overexpression of ARHGAP9 is associated with a good prognosis in hepatocellular carcinoma and bladder cancer. In contrast, we found that high ARHGAP9 expression is related to a poor prognosis in breast cancer[17] and AML[19]. However, the diagnostic and prognostic value and the expression of ARHGAP9 in ccRCC remains unclear. In this report, we conducted a pan-cancer analysis using the GEPIA databases. The results were consistent with those reports that ARHGAP9 is aberrantly expressed in multiple cancers.

Overexpression of ARHGAP9 in ccRCC tissues and its correlation with poor clinicopathologic factors imply that ARHGAP9 is a promising biomarker for the diagnosis of ccRCC. We conducted ROC curve analysis to validate the diagnostic value of ARHGAP9 in ccRCC. The ROC analysis presented an excellent diagnostic value, achieving a sensitivity of 0.958, *specificity* of 0.827 and an *AUC* of 0.950. We further explored the relationship between clinical characteristics and ARHGAP9 expression in ccRCC patients and found that high mRNA expression of ARHGAP9 is significantly associated with advanced TNM stage and lymph node metastases. Since advanced TNM stage and lymph node metastases lead to a lower survival rate, we further conducted Kaplan-Meier plotter and GEPIA2 to validate that up-regulated ARHGAP9 group is associated with a decreased survival rate than the low ARHGAP9 group. These studies indicate that the *ARHGAP9* gene serves as a biomarker in predicting the prognosis of ccRCC patients.

Previous research has shown a correlation between immune cell infiltration into tumors and prognosis. Chen et al[26]. revealed that the ARHGAP family is associated with the prognosis of virous cancers. Hence, we conducted TIMER2.0 and TISIDB to investigate the correlation between tumor infiltrating immune cells and the expression of *ARHGAP9* in ccRCC. We found that ARHGAP9 expression is significantly positive-related to B cells, CD8+ T cells, CD4+ T cells, myeloid derived suppressor cells, and effector memory T cells. Our research provided limited explanations for ARHGAP9 expression and immune infiltration in ccRCC. However, further research needs to be designed to firm up our findings.

In summary, we showed for the first time that ARHGAP9 mRNA is unregulated in ccRcc tissues and the clinicopathological association and function of ARHGAP9 in ccRCC. We also showed that over-expressed ARHGAP9 was positively related to short overall survival, advanced TNM stage, and lymph node metastases in ccRCC patients. Our study suggests ARHGAP9 may be a potential diagnostic and prognostic biomarker in ccRcc and play an important role in immune infiltration of clear cell renal cell carcinoma, and can be used as a therapeutic target for further research.

## Data availability statement

The raw data supporting the conclusions of this article will be made available by the authors, without undue reservation.

## Author contributions

Y.L.X and Y.T designed the study; Y.L.X and C.P collected the data, did the analysis and prepared the manuscript draft. Y.T revised the manuscript. All the authors approved the final proof.

## Data availability statement

The data used to support the findings of this study are available from the corresponding author upon request.

## Data Availability

All relevant data are within the manuscript and its Supporting Information files.
The raw data required to reproduce the above findings are available to download from insert permanent web links.The processed data required to reproduce the above findings are available to download from insert permanent web links.

